# Pleiotropic Loci for Cannabis Use Disorder Severity in Multi-Ancestry High-Risk Populations

**DOI:** 10.1101/2022.11.25.22282743

**Authors:** Qian Peng, Kirk C. Wilhelmsen, Cindy L. Ehlers

**Affiliations:** Department of Neuroscience, The Scripps Research Institute, La Jolla, CA 92037 USA; Department of Neurology, West Virginia University, Morgantown, WV 26506 USA

**Author notes:** Corresponding author, Qian Peng, 3366 North Torrey Pines Court, Suite 200, La Jolla, CA 92037.

## Abstract

Cannabis use disorder (CUD) is common and has in part a genetic basis. The risk factors underlying its development likely involve multiple genes that are polygenetic and interact with each other and the environment to ultimately lead to the disorder. Co-morbidity and genetic correlations have been identified between CUD and other disorders and traits in select populations primarily of European descent. If two or more traits, such as CUD and another disorder, are affected by the same genetic locus, they are said to be pleiotropic. The present study aimed to identify specific pleiotropic loci for the severity level of CUD in three high-risk population cohorts: American Indians (AI), Mexican Americans (MA), and European Americans (EA). Using a previously developed computational method based on a machine learning technique, we leveraged the entire GWAS catalog and identified 114, 119, and 165 potentially pleiotropic variants for CUD severity in AI, MA, and EA respectively. Ten pleiotropic loci were shared between the cohorts although the exact variants from each cohort differed. While majority of the pleiotropic genes were distinct in each cohort, they converged on numerous enriched biological pathways. The gene ontology terms associated with the pleiotropic genes were predominately related to synaptic functions and neurodevelopment. Notable pathways included Wnt/β-catenin signaling, lipoprotein assembly, response to UV radiation, and components of the complement system. The pleiotropic genes were the most significantly differentially expressed in frontal cortex and coronary artery, up-regulated in adipose tissue, and down-regulated in testis, prostate, and ovary. They were significantly up-regulated in most brain tissues but were down-regulated in the cerebellum and hypothalamus. Our study is the first to attempt a large-scale pleiotropy detection scan for CUD severity. Our findings suggest that the different population cohorts may have distinct genetic factors for CUD, however they share pleiotropic genes from underlying pathways related to Alzheimer’s disease, neuroplasticity, immune response, and reproductive endocrine systems.

## INTRODUCTION

Cannabis is the most commonly used psychotropic substance in the U.S., after alcohol. The prevalence of use appears to be increasing, likely due to legalization in many states and decreasing perception of harm, especially among young people [1, 2]. About one-fifth of lifetime cannabis users meet criteria for DSM-5 use disorder (CUD), and nearly a quarter are diagnosed as severe [3]. Twin and family studies have consistently found that cannabis use and use disorders appear to in part have a genetic basis (for review, see [4]). Twin samples have found estimates of heritability that range from 0.45 to 0.78 [5-12], suggesting that studies aimed at identifying genes that contribute to cannabis involvement may be warranted.

A recent meta-analysis of “cannabis use” provides evidence to suggest that the genetic basis of cannabis use is polygenetic, that is being influenced by multiple genes and also the environment. In those sets of analyses 96 genetic variants were identified that met significance threshold (p< 1.0 × 10^−7^) [13]. Cannabis use may be influenced by a number of genes that are difficult to detect because they may each have a small effect on the broad clinical phenotypes. However, the genes influencing *cannabis use disorders* might be detected if they have a major effect on more narrowly defined phenotypes that more closely index the biological processes associated with severe addiction. Additionally, cannabis use disorders are often comorbid with other health issues especially mental health problems, further complicating the genetic analyses [14]. Both CUD and other mental health disorders have been shown to have a significant genetic component to their etiology [13, 15, 16]. However, documenting comorbidity between disorders does not necessarily imply a common etiological pathway or a causal connection between them. Behavioral genetics studies have an advantage in being one of the most powerful methods for determining whether the comorbidity among psychopathological conditions is due to common pathologies and/or etiologies associated with both disorders.

In addition to polygenicity, cannabis phenotypes can also be impacted by pleiotropy. Two or more traits are formally said to be “pleiotropic” if they are affected by the same genetic locus [17]. Given the polygenic nature of cannabis use and use disorders and their genetic correlations to other diseases [18], these pleiotropies are also expected to be polygenic. For instance, one study found a pleiotropic linkage peak for cannabis use and major depression in a Mexican American population [19]. Additionally, a recent study based on large scale GWAS meta-analyses identified 15 loci for multiple substance use including cannabis and other psychopathology, and five pleiotropic loci specific for lifetime cannabis use and psychiatric disorders [20]. Few studies however have identified the *specific* pleiotropic loci for CUD and other co-morbid disorders. One attempt to uncover shared genetic variation between cannabis dependence and five other psychiatric disorders identified *CSMD1* as a candidate gene that may affect the risk for both cannabis dependence and schizophrenia [21].

While cannabis use and CUD are genetically correlated to some extent (*r*=0.5), there is also clear distinction between the genetic liability to “cannabis use” and “cannabis use disorder” [18]. The pleiotropic landscape of CUD with other diseases therefore also likely differs from that of cannabis use. In addition, while the large GWAS studies of CUD to date have included multiple ancestries, the findings by these studies are still predominately from the populations of European descent, driven by the much larger sample sizes of these populations compared to populations of other ancestries in the studies [18, 22]. Prevalence of CUD and degrees of severity however vary significantly across ethnic groups and populations. For instance, epidemiology studies have shown that American Indians have significantly increased rates of CUD, especially in the severe cases [3, 23, 24]. The differences are likely due to both environmental and genetic factors.

While it is likely that multiple factors contribute to the comorbidity of complex diseases such as CUD, the present study attempts to address three related fundamental questions: 1) what shared genetic factors may underly CUD and comorbid disorders; 2) what other diseases and phenotypes may be linked to CUD through genetics; and 3) are there population differences in genetic factors associated with CUD. In the present study, we focus on pleiotropic detection for a quantitative cannabis use disorder phenotype that indexes the *severity of the disorder* in three independent population cohorts including American Indians (AI), Mexican Americans (MA), and European Americans (EA). All three cohorts had elevated rates of CUD of varying degrees compared to the general populations: 39% in AI, 27% in MA, and 21% in EA respectively. Previous studies on the EA cohort have found cannabis use and dependence heritable [25], and identified two low-frequency variant loci associated with cannabis dependence through a meta-analysis of the EA and AI [26]. In the AI cohort, a cannabis dependence cluster phenotype was found to be heritable and significant evidence for genetic linkage analyses was found on chromosome 16 and 19 [27]. To investigate the degree of CUD severity as well as increase statistical power for detection, we used a phenotype that was derived from the clinical course of CUD to quantitate the CUD severity. We previously described a flexible computational framework to explore potential pleiotropy for complex disorders, as illustrated in Figure 1. The method was designed especially with understudied admixed populations in mind where few genetic studies were available for pleiotropic analyses [28]. We herein applied the method to the pleiotropic detection for CUD severity in the three high-risk population cohorts and investigated their distinct and shared pleiotropic traits and pathways.

**Figure 1.**
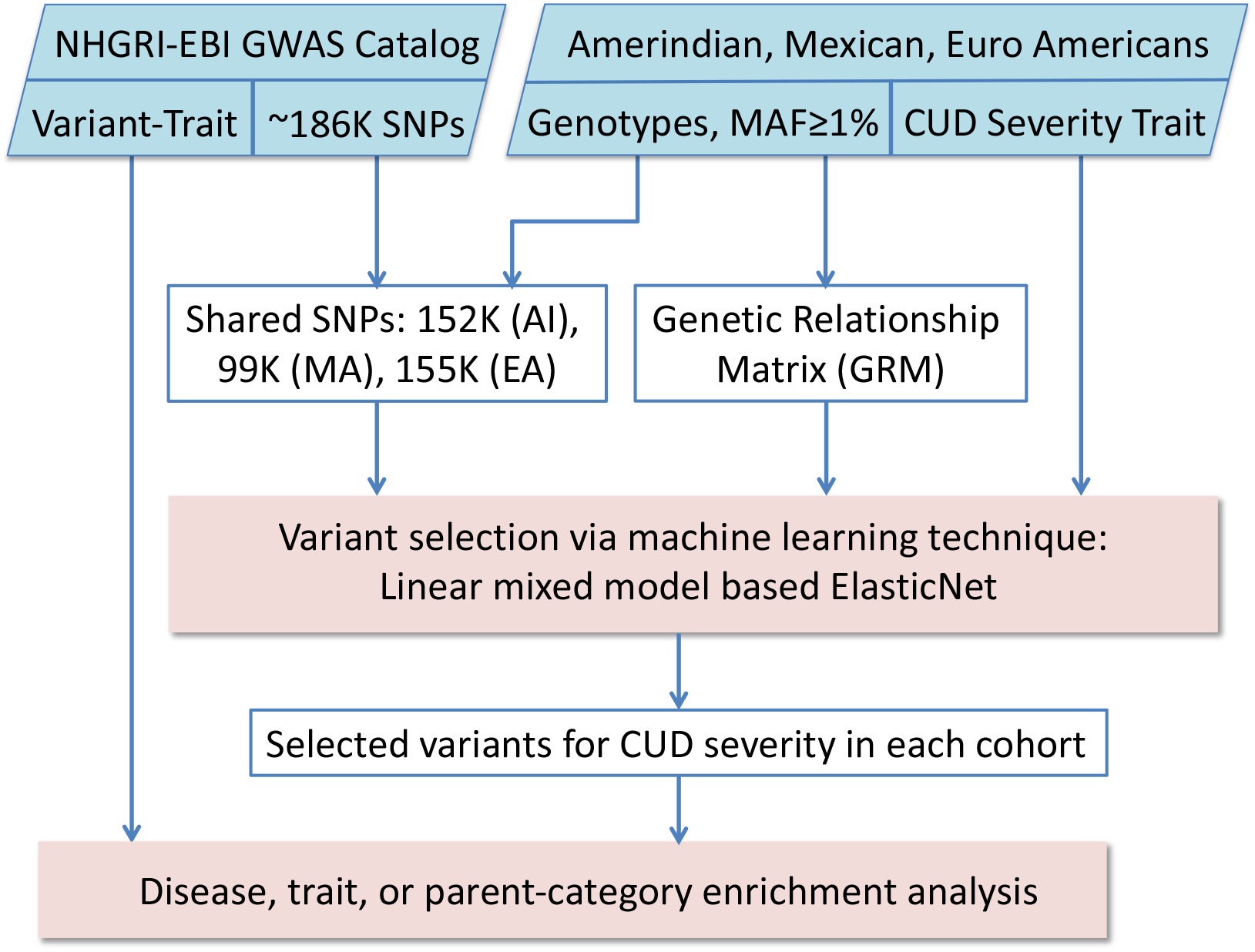
Illustration of the CUD pleiotropy analysis pipeline.

## MATERIALS AND METHODS

### Participants

We investigated three independent populations including an American Indian (AI) cohort of 742 participants from extended pedigrees, a Mexican American (MA) cohort of 547 participants from primarily second-generation Mexican American young adults, and a Euro-American (EA) cohort of 1711 participants from the San Francisco Family Alcohol Study. We refer to the first cohort as AI, the second as MA, and the third as EA. The population characteristics and the recruitment procedures of the three cohorts have been previously described [29-36]. Their demographics are characterized in Table S1. The protocol of the study of the American Indian cohort was approved by the Scripps Research Institute Institutional Review Board (TSRI-IRB) and presented to Indian Health Council, a tribal review group overseeing health issues for the reservations where recruitments took place. The protocol of the study of the MA cohort was also approved by TSRI-IRB. The protocol for collection of participants in the EA cohort was approved by the University of California San Francisco (UCSF) Committee for the Protection of the Rights of Human Subject while the recruitment took place. Subsequently, the University of North Carolina, at Chapel Hill IRB approved the data analysis plan. Written informed consent was obtained from each participant after study procedures had been fully explained. Participants were compensated for their time spent in the study.

### Phenotypes and genotypes

All participants were deep-phenotyped using an instrument called Semi-Structured Assessment for the Genetics of Alcoholism (SSAGA) [37, 38]. The CUD severity is a derived phenotype that quantifies the progression and severity of the use disorder. We extended measures of the clinical course of alcoholism originally described by Schuckit and colleagues [39] to quantitate life events during the clinical course of other substance use and psychiatric disorders [28, 30, 40, 41]. The measures were based on the relative order of the appearance of major “substance-related life events”. Table S2 lists 20 cannabis-related life events in the clinical course of CUD. The order of the events within each population cohort was determined by the mean age of occurrence of these events if available, with the first event occurring the earliest and the last event (the 20^th^) the latest in the lifetime. The exact orders of events thus may vary between different population cohorts. We then developed a metric that quantitates the severity level of CUD progression based on the sequential occurrence of the events. The CUD clinical course life events were each given a severity weight as shown in Table S2: events 1-7 each have weight 1, events 8-14 have weight 2, and 15-20 have weight 3. The metric gives larger weights to the more severe events that occur later in the clinical course of CUD and are more associated with severe use disorder [40, 42]. Finally, the CUD severity phenotype was defined as the total sum of the severity weights of the life events that occurred in an individual [42]. The distributions of the phenotype for each cohort and their relations to the DSM-5 CUD diagnoses are shown in Figure S1. The quantitative CUD severity phenotype is in high accordance with DSM-5 diagnoses, with the Spearman’s rank correlation ρ between the CUD severity and the CUD diagnosis (no, mild, moderate, and severe) being 0.90, 0.93, and 0.89 for AI, MA, and EA respectively. We winsorized CUD severity level at 5% at each tail for subsequent analysis.

The AI and EA participants had low-coverage whole genome sequencing on blood-derived DNAs using the same pipeline [43]. The MA participants had exome data [44]. Further details can be found in SI methods.

### Variant-trait association catalog

We only considered single nucleotide polymorphisms (SNPs) in this study thus we will use terms SNP and variant interchangeably. We downloaded the GWAS catalog release v1.0.2 on 2021-12-21 from NHGRI-EBI [45]. This release contained 325,538 SNP-trait associations from 5,527 publications. Genes to which a SNP is mapped were also reported for each association. Of the 186,481 unique variants reported in the catalog, 152,074 were found in the AI cohort, 99,386 in the MA cohort, and 154,580 in the EA cohort, respectively. We referred to these variants from each cohort as the *candidate variant set* for that cohort. The catalog had total of 3,061 uniquely mapped traits (terms from the experimental factor ontology (EFO) to which GWAS reported traits were mapped) that were classified into 17 parent categories [46].

### Pleiotropic variant selection

We updated a previously described method to identify a set of variants for each cohort from its candidate variant set to be associated with CUD severity [28]. Briefly, a machine learning technique called elastic net—a balanced regularization between lasso and ridge penalties—was used for variant selection [47]. The genetic relationship matrix (GRM) was incorporated into the model to control for both population structures and potential relatedness. Age, age-squared, and sex were included as covariates. The GRM for each cohort was estimated with GCTA [48]. The *R* package *glmnet* was used to solve the elastic net optimization. The elastic net penalty α that bridges the gap between lasso and ridge regressions was set to 0.5. The regularization parameter λ that controls for the overall strength of the penalty, thus the number of variants that are eventually selected, was determined through cross validations. We did 10 rounds of 10-fold cross validation and chose the minimum λ. The variants that were selected by this process are referred to as *pleiotropic variants* for CUD severity in each cohort.

### Pleiotropic trait & disease enrichment and functional analysis

GWAS catalog reported variants along with their mapped genes. We refer to the genes that the pleiotropic variants are mapped to as *pleiotropic genes*. To determine whether a trait or disease is overrepresented in the set of pleiotropic variants, we defined trait enrichment as the ratio of the proportion of the pleiotropic genes that are associated with a trait in the GWAS catalog to the proportion of the genes which the candidate variants are mapped to that are associated with the same trait. The significance of the enrichment was determined through 10,000 permutation tests.

We combined pleiotropic genes from the three cohorts and subjected them to functional analysis. Functional enrichment analyses were performed using GENE2FUNC in FUMA version 1.4.1, where enriched biological functions or pathways were extracted by testing against gene sets from the molecular signatures database (MsigDB) [49] and WikiPathways [50] using hypergeometric tests [51]. The tissue-specific differential gene expression test was conducted for the combined pleiotropic gene set against all genes across genomes that exhibited significantly increased or decreased expression levels in a certain tissue sample compared to all other samples. The analysis utilized tissue-specific transcriptome data across 54 tissue types from GTEx v8 [52].

### Shared pleiotropic genes and loci

If a pleiotropic gene appeared in more than one cohort, we called it a *shared pleiotropic gene*. Pleiotropic variants in linkage disequilibrium (LD) with each other in the same cohort were considered as one pleiotropic locus. If two loci across two cohorts overlapped in their genomic positions, they were also considered as a *shared pleiotropic locus* [28].

## RESULTS

### Pleiotropic variants and enriched pleiotropic traits for CUD severity

One hundred and fourteen (114) SNPs in the GWAS catalog were selected as pleiotropic variants for CUD severity in the AI cohort, while 119 and 165 SNPs were selected in the MA and EA cohorts respectively. All identified pleiotropic variants are listed in Table S3. Figure 2 illustrates the enriched pleiotropic traits with a minimum of three SNPs selected for the trait for CUD severity in each cohort. Alzheimer’s disease (AD) is the most enriched in AI with four pleiotropic SNPs (enrichment=7.2, *p*=0.003) followed by PHF-tau measurement which is an AD biomarker (enrich=5.6, *p*=0.0006). The mapped genes for Alzheimer’s disease are *ABCA1, ACE, APOE-APOC1* and *EPHA1* (see Table S3). The most enriched trait in MA is testosterone measurement (enrich=4.3, *p*=0.033), while brain volume (enrich=4.5, *p*=0.028) is the most enriched in EA. MA and EA each have an enriched anthropometric trait, body weight in MA (enrich=4.2, *p*=0.037) and BMI-adjusted waist-hip ratio in EA (enrich=3.6, *p*=0.0026). Mean platelet volume is enriched in both AI and MA. The complete list of enriched traits and diseases (*p*<0.05) including traits that were mapped to fewer than 3 variants are given in Table S4.

**Figure 2.**
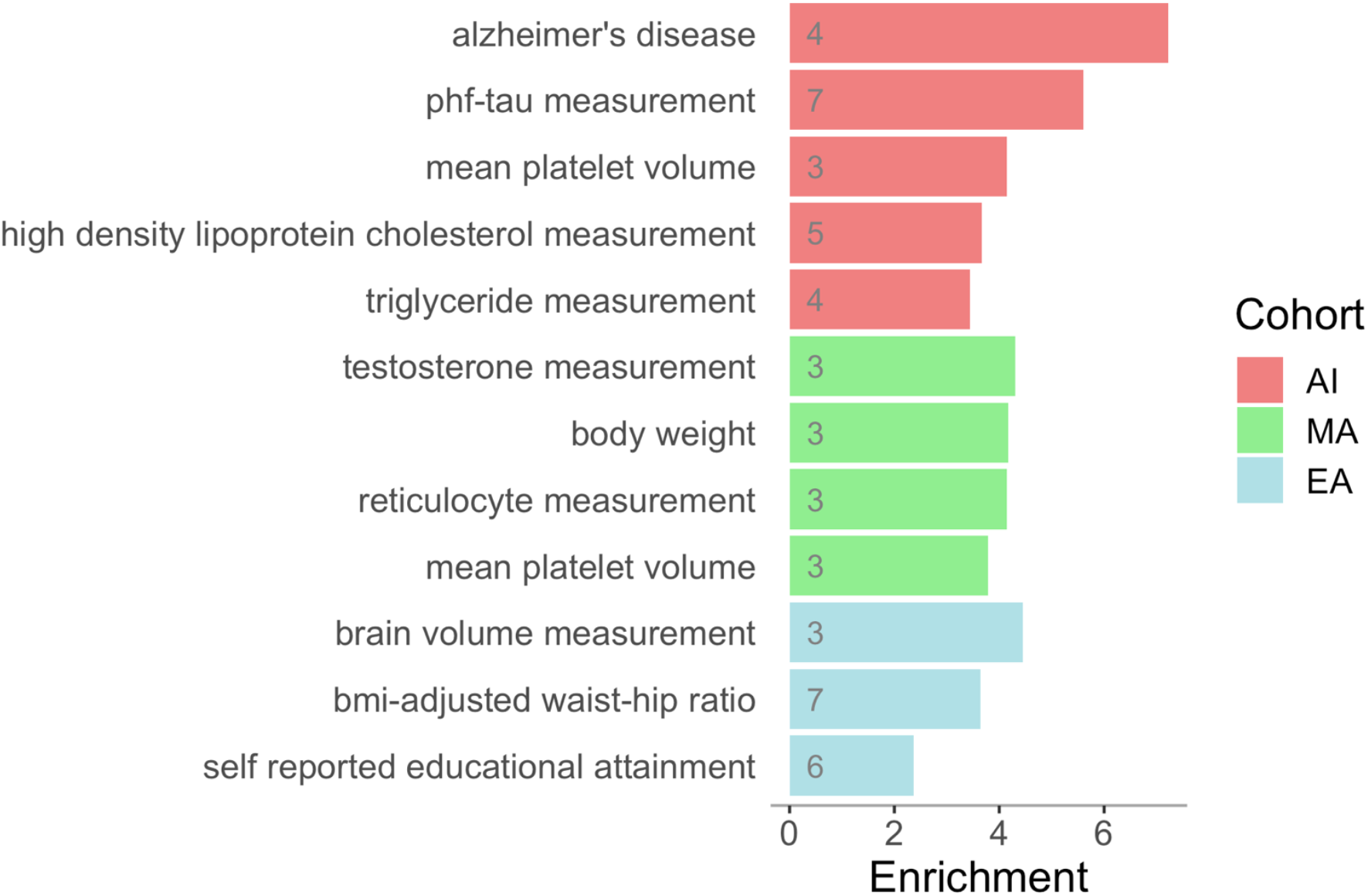
Significantly enriched pleiotropic traits and diseases for CUD severity in the American Indian (AI), Mexican American (MA), and European American (EA) cohorts. The number in each enrichment bar indicates the number of SNPs selected from the GWAS catalog for the respective trait (see Tables S3 and S4 for complete details).

In addition, two variants in MA (mapped to genes and intergenic regions near *PLSCR5* and *SLC4A7*) and three variants in EA (mapped to genes *CELF2, TENM2*, or near *LINC01924*) were identified for smoking status or cigarettes per day. Two variants from AI and one from EA were selected for drinking behaviors. One variant each from AI and EA were identified for “substance abuse”. One variant from AI (near *CDH18*) and three variants from MA (*TUBGCP6, OSTM1, KCNK13-PSMC1*) were selected for unipolar depression. One variant from AI and two from EA (*MAP1B*, intergenic near *ABHD5*) were selected for ADHD. One variant from AI, three variants from MA, and four variants from EA were identified for schizophrenia. Four variants from MA and six from EA were selected for educational attainment. Every cohort had pleiotropic variants identified for lipid or lipoprotein measurements: six in AI (mapped to *ABCA1, APOE-APOC1, CPNE6, EPHA1, INPP4B, and LINC02644-ZNF438*), six in MA (mapped onto or near *CLPTM1, DPP3, BSND-PCSK9, PDGFD*, and *NOTCH1*), and eight in EA (mapped to genes *DNASE2B, RIPK4, PPP6R2*, or intergenic near *TOX3, FTLP17, MED27-NTNG2, PDHX-CD44*, and *NUFIP2-RPL35AP35*). Eight variants from AI, 18 from MA, and 24 from EA were selected for anthropometric traits such as BMI and waist-hip ratio (see Tables S3A-C).

### Pleiotropic genes and loci shared by cohorts

Pleiotropic variants selected by each cohort were unique. However, as shown in Table 1, there were several genes and loci identified by more than one cohort even though the exact variants differed. Genes *DAB1* and *PTPRG* were selected in both MA and EA. *DAB1* encodes a reelin adaptor protein that is essential for positioning migrating neurons in the developing brain and during adult neurogenesis [53]. *PTPRG* is a protein tyrosine phosphatase receptor gene that is also associated with schizoaffective disorder. Genes *CDH18, ERBB4, SCGB1A1* and a LncRNA *DLEU1* were selected in both AI and EA. Cadherin-18 family gene *CDH18* mediates calcium-dependent cell-cell adhesion. *SCGB1A1* encodes uteroglobin, a founding member of the secretoglobin family of small secreted proteins. *ERBB4* is a neuregulin receptor and has been implicated in schizophrenia. Neuregulins and their receptor ErbB4 play important roles in the development of the central nervous system, specifically in the differentiation of GABAergic interneurons. In addition to schizophrenia [54], *ERBB4* has also been associated with smoking behaviors [55], cognitive performance [56], neuroticism [57], psychosis [58] and so on. The only locus selected by all three cohorts is an ephrin receptor gene *EPHA1* and EPHA1 antisense RNA 1 *EPHA1-AS1*. Its associated traits were either Alzheimer’s disease or a biomarker measurement for AD. *EPHA1* has been implicated in developmental events especially in the nervous system. Two additional shared loci between AI and EA, and one shared locus between MA and EA were identified based on LD measures (Table 1). Each locus consisted of two or three genes.

**Table 1.** Pleiotropic genes and loci from GWAS catalog identified in multiple cohorts for CUD severity.

| Gene/Locus | dbSNP | Position (hg19) | Pleiotropic GWAS Trait | AI | MA | EA |
| --- | --- | --- | --- | --- | --- | --- |
| *DAB1* | rs264038 | 1:57669814 | household income |  | x |  |
|  | rs536250 | 1:57570761 | adolescent idiopathic scoliosis |  |  | x |
|  | rs1884486 | 1:58330801 | waist circumference |  |  | x |
| *ERBB4* | rs7564590 | 2:213387900 | polycystic ovary syndrome | x |  |  |
|  | rs59779307 | 2:212642775 | household income |  |  | x |
| *PTPRG* | rs652889 | 3:61794054 | qt interval |  | x |  |
|  | rs9825906 | 3:61993704 | body height |  |  | x |
| *CDH18* | rs349475 | 5:19440168 | unipolar depression | x |  |  |
|  | rs1498103 | 5:19839503 | response to TNF antagonist |  |  | x |
| *EPHA1* | rs10808026 | 7:143099133 | Alzheimer's disease, HDL cholesterol | x |  |  |
| *EPHA1-AS1* | rs75045569 | 7:143109208 | Alzheimer's disease |  |  | x |
|  | rs17382348 | 7:143204912 | PHF-tau measurement |  | x |  |
| *SCGB1A1* | rs3018614 | 11:62174721 | BMI-adjusted waist-hip ratio |  |  | x |
|  | rs2509973 | 11:62179371 | BMI-adjusted waist-hip ratio |  |  | x |
|  | rs17145884 | 11:62200176 | serum gamma-glutamyl transferase (GGT), serum alanine aminotransferase (ALT) | x |  |  |
| *DLEU1* | rs797487 | 13:51224309 | adolescent idiopathic scoliosis | x |  |  |
|  | rs12429206 | 13:51446114 | prostate specific antigen measurement |  |  | x |
| *ZNF192P2-TOB2P1* | rs1150687 | 6:28162469 | age at onset, myopia, refractive error | x |  |  |
| *NKAPL* | rs1635 | 6:28227604 | schizophrenia |  |  | x |
| *DPP3* | rs148812168 | 11:66251451 | LDL cholesterol measurement |  | x |  |
| *RHOD* | rs11602157 | 11:66827319 | BMI-adjusted hip circumference |  |  | x |
|  | rs3923203 | 11:66835194 | BMI-adjusted waist circumference |  |  | x |
| *ASCC2-MTMR3* | rs5763555 | 22:30251180 | heel bone mineral density |  |  | x |
| *HORMAD2* | rs4820829 | 22:30524248 | testosterone measurement | x |  |  |

### Enriched pathways in pleiotropic genes for CUD severity

The selected pleiotropic variants mapped to total of 439 unique genes—referred to as pleiotropic genes—in the three cohorts combined (137 in AI, 122 in MA, and 190 in EA). The pleiotropic genes from the three cohorts converged onto numerous enriched functional groups and pathways. Figure 3 illustrates the enriched pathways [50, 59-61] and hallmark gene sets from MSigDB [62], and gene ontology sets [63]. A more comprehensive list of enriched gene sets is given in Table S5.

**Figure 3.**
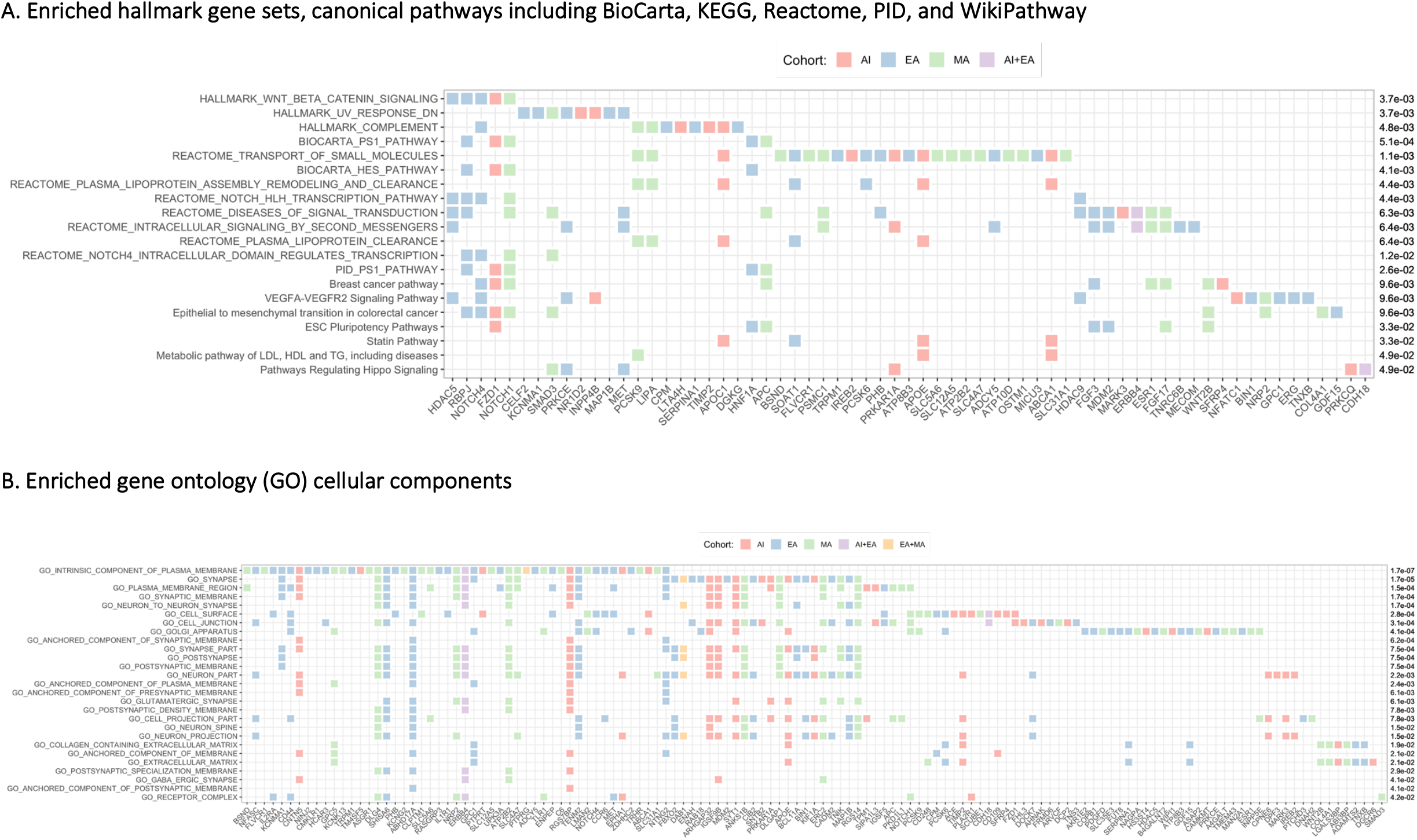
Significantly enriched pathways and gene sets of pleiotropic genes selected for CUD severity in the American Indian (AI), Mexican American (MA), and European American (EA) cohorts.

Three hallmark gene sets were significantly enriched. They were genes up-regulated by activation of Wnt signaling through accumulation of beta catenin CTNNB1, genes down-regulated in response to ultraviolet (UV) exposure, and genes encoding components of the complement system. Presenilin-1 action in Notch and Wnt signaling was the most enriched pathway, followed by transport of small molecules. Lipoprotein functions, breast cancer and colorectal cancer related pathways, and a vascular endothelial growth factor (VEGF) signaling pathway were also among the enriched pathways.

Of the gene ontology (GO) cellular components, gene sets related to synapse, cell junction, Golgi complex, and extracellular matrix (ECM) were found to be significantly enriched (Figure 3B). Neuron differentiation, cell cycle, adhesion, inflammatory response, lipid function, locomotion, ion transport, and homeostasis were among the enriched GO biological processes. There were also numerous enriched transcription factor (TF) targets and microRNA target gene sets, and over one hundred enriched transcriptional immunologic signatures [64] (Table S5).

### Tissue-specific differential expression in pleiotropic genes

Figure S2 illustrates the enrichment in tissue-specific differentially expressed gene sets. The identified pleiotropic genes are said to be significantly enriched in a tissue if they consisted of an overrepresentation of genes that were significantly more (up regulated) or less (down regulated) expressed in the given tissue compared to all other tissues [51]. The pleiotropic genes were the most significantly differentially expressed in coronary artery and frontal cortex (adjusted *p*=3.2E-5, 5.5E-5; subsequent numbers in parenthesis are adjusted *p* values). They were the most significantly up regulated in adipose tissue (1.7E-4), and down regulated in testis (1.4E-4), prostate (1.7E-4) and ovary (2.7E-4), followed by adrenal gland (8.0E-4) and esophagus mucosa (2.3E-3). Pituitary was also down regulated (5.6E-3). Almost all brain tissues tested were significantly enriched in differentially expressed pleiotropic genes. Among the brain tissues, hippocampus (4.5E-3), substantia nigra (1.4E-3), cortex (5.7E-3), nucleus accumbens (0.015), putamen (0.020), frontal cortex (0.035), and amygdala (0.046) had significantly up regulated pleiotropic genes; Cerebellum (2.4E-3) and hypothalamus (0.034) had significantly down regulated pleiotropic genes; Caudate nucleus (0.012) and anterior cingulate cortex (0.022) had significantly differentially expressed pleiotropic genes.

## DISCUSSION

Over the last two decades cannabis use and use disorders have increased globally [65]. Cannabis use among youth, especially in minority communities like American Indians and Mexican Americans is particularly problematic because early use had been shown to lead to an increased risk for use disorders [66, 67] as well as other symptoms such as depression in the populations investigated [68]. Cannabis use and use disorder risk is in part genetic and identifying novel variant-trait associations through genome-wide approaches can potentially lead to the identification of select biological mechanisms related to the disorder that could inform interventions.

The present study addressed three related fundamental questions in cannabis genetics research: 1) what shared genetic factors may underly CUD and comorbid disorders, 2) what other diseases and phenotypes may be linked to CUD through genetics, and 3) are there population differences in genetic factors associated with CUD. In the set of analyses conducted for the present report, over 100 potentially pleiotropic variants for CUD severity were identified in each of the three population cohorts that had elevated rates of CUD, including American Indians, Mexican Americans, and European Americans. Although the variants and most genes from each cohort were distinct, the genes to which these variants were mapped have converged onto numerous enriched biological pathways. There were also several genes and loci shared between the cohorts. The gene ontology terms associated with the pleiotropic genes were predominately related to synaptic functions and neurodevelopment. Notable pathways included Wnt/β-catenin signaling, lipoprotein assembly, response to UV radiation, and components of the complement system. The Wnt/β-catenin pathway mainly controls cell proliferation and regulates cortical size [69, 70]. The Wnt/β-catenin signaling is also associated with oxidative stress, inflammation, and the glutamatergic pathway. Animal studies have shown that this pathway is critical in drug neuroadaptation by inducing behavioral sensitization as indexed by β-catenin levels: β-catenin decreased in certain brain regions in animals that showed drug-induced sensitization [71]. Additionally, cannabidiol (CBD), a less psychoactive cannabis component, has been shown to increase the activity of the WNT/β-catenin pathway [72]. CBD also protects against the harmful effects of UV radiation by decreasing the expression of proinflammatory proteins and the proteins involved in de novo protein biosynthesis, which are elevated in UV-irradiated cells [73].

### Cognitive functions and psychiatric disorders

Since cannabinoid receptor 1 (CB1) is highly expressed within the central nervous system and endocannabinoids are key modulators of synaptic function [74], it is not surprising that synaptic functions and neuron structures were significantly overrepresented in the pleiotropic genes identified for CUD severity. Pleiotropic genes were also significantly differentially expressed in almost all brain tissues, mostly up-regulated with the exceptions of cerebellum and hypothalamus where the pleiotropic genes were significantly down-regulated. Brain volume was the most enriched pleiotropic trait for CUD severity in EA. While educational attainment was an enriched pleiotropic trait for EA, associated variants were also found in MA. Variants associated with smoking, drinking or other substance use were found for CUD in each cohort. Both AI and MA identified pleiotropic variants for depression, and in AI and EA variants were identified for ADHD. All three cohorts had pleiotropic variants associated with schizophrenia.

Alzheimer’s disease (AD) and AD markers were the most enriched pleiotropic traits for the AI cohort. Interestingly, the only pleiotropic gene locus for CUD severity that was found in all three cohorts is an ephrin receptor gene *EPHA1* and *EPHA1-AS1*. The traits associated with the variants on this locus were Alzheimer’s disease, AD biomarker, and HDL cholesterol. EphA receptors are mediators for axion guidance, and *EPHA1* has been implicated in developmental events especially in the nervous system. In addition, presenilin-1 (PS1) signaling pathway was significantly associated with the combined set of pleiotropic genes selected by the three cohorts. PS1 is linked to gamma secretase activity that cleaves amyloid precursor protein [75] and has been implicated in Alzheimer’s disease [76]. Wnt pathway disruption by β-amyloid (Aβ) peptide also represents a pivotal event in the neuronal apoptosis occurring in AD [77]. Additionally, animal studies have implicated blocking of endocannabinoids in the early pathology of AD. β-Amyloid blocks synaptic plasticity via loss of cannabinoid-mediated disinhibition [78].

Pleiotropic genes were also found to be significantly associated with lipoprotein pathways. HDL and triglyceride levels were enriched pleiotropic traits in AI. Body weight and waist-hip ratio were enriched for MA and EA respectively. In the 10 loci shared among the cohorts, four had variants associated with a cholesterol level or a body measurement. Every cohort had pleiotropic variants identified for lipid or lipoprotein measurements and for body measurements such as BMI and waist-hip ratio.

### Immune response

Components of the complement system—part of the innate immune system—was significantly associated with pleiotropic genes. Inflammatory response and lipid function were among significantly enriched GO biological processes. There were also over a hundred transcriptional immunologic signatures [64] enriched in the pleiotropic genes. Additionally, *SCGB1A1* was identified in both AI and EA. The gene encodes uteroglobin, a multifunctional protein with anti-inflammatory and immunomodulatory properties [79]. Immune red blood cell reticulocyte count was enriched pleiotropic trait in EA. These findings in general are consistent with the immune modulation functions of the cannabinoid system. Cannabinoid receptor 2 (CB2) is largely found in the immune cells. Strong evidence supports that CBD suppresses the immune activity and inflammatory response [80]. Δ-9-tetrahydrocannabinol (THC), the main psychoactive component of cannabis, is also known to alter the function of immune cells and exert immune depressive effects [81].

### Reproductive and endocrine systems

The selected pleiotropic genes were most significantly down regulated in testis, prostate, and ovarian tissues. They were also significantly down regulated in adrenal gland, pituitary, hypothalamus, and vagina, and differentially expressed in the mammary tissue. Level of testosterone was the most enriched pleiotropic trait for CUD severity in MA. One locus identified in both AI and EA contained a variant that was associated with testosterone level. Additionally, several variants were selected for carcinoma or tumors related to the reproductive and endocrine systems, including four variants (*GDF7*) for prostate cancer in MA, one each in MA and EA for uterine fibroid, one in AI for ovarian cancer, one in AI for thyroid cancer, two in AI (*SHLD1, CDYL2*) and one in EA (*TACC2*) for breast cancer. A breast cancer pathway, several breast cancer related gene sets, and one prostate cancer related gene set were significantly enriched (Fig 3A, Table S5). Cannabinoid receptors are present throughout the body including hypothalamus, pituitary, ovary, endometrium, testes, and spermatozoa. Interactions of cannabinoids with hypothalamic–pituitary–gonadal axis hormones are well documented [82]. Research to date have suggested that cannabis use may affect some of the central reproductive process [83]. Although the overall relations between cannabis use and cancer risk remain unclear, there are evidence suggesting that long time cannabis use may increase the risk for testicular cancer [84].

In addition to breast cancer and prostate cancer, a colorectal cancer-related pathway was also enriched. Endocannabinoid signaling system modules many pathways that are often dysregulated in cancer, such as cell proliferation, motility, and survival. There are evidences of increased CB1 expression in prostate cancer and colon cancer, and increased CB2 expression in breast cancer [85].

Several pleiotropic genes for CUD severity identified in this study were previously associated with cannabis use or dependence. Variant rs11576941 associated with eosinophil count on the fatty acid amide hydrolase gene (*FAAH*) was selected in MA. *FAAH* is involved in endocannabinoid metabolism and was a candidate gene previously identified for cannabis use and dependence [86] including the MA population under study [87]. Rs2724992 on the CUB and sushi multiple domains 1 (*CSMD1*) gene was selected in AI. Different SNPs on *CSMD1* have previously been associated with cannabis dependence [21] and schizophrenia [88]. *CSMD1* is highly expressed in the growth cones of developing central nervous system (CNS) neurons suggesting that it may act as an important regulator of complement activation and inflammation in the developing CNS [89]. Rs66579625 associated with body height on *INTS7* was found in MA. Two different SNPs on *INTS7* have been associated with DSM-IV cannabis dependence criteria count [21]. Rs3943782 on *CADM2* associated with risk-taking behavior was selected in EA. A different SNP on *CADM2* was associated with lifetime cannabis use [90]. Rs6907357 was selected in MA. The variant was reported to interact with gene *FAM19A5* (a.k.a *TAFA5*). A SNP on *FAM19A5* has been associated with CUD [22]. *FAM19A5* encodes a neurokine that plays a crucial role in depressive-like and spatial memory-related behaviors [91].

### Methodological considerations and future directions

Our findings should be interpreted in light of the study design along with limitations. We used the term pleiotropy broadly and our method for detection was through cross-trait association. As a result, our findings did not distinguish between horizontal (biological) pleiotropy, where causal variants for different traits are on the same locus, and vertical (mediated) pleiotropy, where a genetic factor affects a trait indirectly through another trait [92]. In addition, variants associated with a phenotype may not be the causal variants, thus some of our findings may be the results of one variant tagging different causal variants for different traits through LD, or one variant being linked to a disease by indexing other traits. Statistical fine mapping and Mendelian randomization will be needed to further determine the pleiotropic scenarios for each potentially pleiotropic variant we identified, followed by experimental studies to confirm the true biological pleiotropy. GWAS catalog associations are currently dominated by studies of European descent. Our pleiotropic findings for CUD severity therefore might also be biased toward genetic factors associated with diseases in European ancestry. However, we were still able to identify many enriched pathways from the pleiotropic genes contributed by all three cohorts of different ancestries. It should also be noted that the MA cohort only had exome data thus precluding most of the intergenic regions, although the functional analysis focused primarily on genes and their associated pathways. Finally, we acknowledge that we were limited by sample sizes in all three population cohorts. We did, however, identify 10 pleiotropic loci that were shared among the cohorts for CUD severity. This was achieved by focusing on high-risk populations, deriving a well-defined quantitative trait from deep-phenotyped study cohorts, leveraging a large public database, and employing a computational approach for dimension reduction. Note that most of the pleiotropic variants identified in this study would not be associated with CUD phenotypes at a genome-wide significant level if tested individually in GWAS.

In conclusion, we used a previously developed computational method to carry out the first large-scale pleiotropic detection for CUD severity in American Indians, Mexican Americans, and European Americans. Our findings suggest that while each population cohort had distinct variants and genes for CUD and pleiotropic traits, the potentially pleiotropic genes from different cohorts also converged on enriched pathways related to Alzheimer’s disease, neuroplasticity, immune response, and reproductive and endocrine systems.

## Data Availability

The data from the European American cohort under study is available in dbGaP, study accession phs001458.v1.p1. The data from the American Indian cohort under study cannot be shared publicly in accordance with the wishes of the tribes. All other data produced in the present study are available upon reasonable request to the authors.

## ACKNOWLEDGEMENTS

We would like to acknowledge and thank all our research participants, and the following people for their roles in 1) the genotyping effort: Scott Chasse, Piotr Mieczkowski, Ewa Patrycja Malc, Joshua Sailsbery, Phil Owens, and Chris Bizon; and 2) recruiting participants, and collection and preparation of the clinical data: David Gilder, Corinne Kim, Evie Phillips, Gina Stouffer, Susan Lopez, Linda Corey, Greta Berg, Phillip Lau, and Derek Wills.

This work was supported by the National Institutes of Health (NIH): National Institute on Drug Abuse (NIDA) DP1 DA054373 to QP; R01 DA030976 to CLE. National Institute on Alcohol Abuse and Alcoholism (NIAAA) K25 AA025095 to QP; R01 AA027316 to CLE. NIAAA and NIDA had no further role in the study design; in the collection, analysis, and interpretation of data; in the writing of the report; or in the decision to submit the article for publication.

## CONFLICT OF INTEREST

The authors declare no conflict of interest.

## Notes

### Competing Interest Statement

The authors have declared no competing interest.

### Author Declarations

IRB of The Scripps Research Institute gave ethical approval for this work. Committee for the Protection of the Rights of Human Subject of the University of California San Francisco gave ethical approval for this work. IRB of the University of North Carolina, at Chapel Hill gave ethical approval for this work.

